# Human amyotrophic lateral sclerosis/motor neuron disease: The disease-associated microglial pathway is upregulated and APOE ε2 is protective

**DOI:** 10.1101/2025.02.07.25321850

**Authors:** Bridget A Ashford, Julie E Simpson, Charlotte Dawson, Delphine Boche, Johnathan Cooper-Knock, Paul R Heath, Daniel Fillingham, Charlie Appleby-Mallinder, Wenbin Wei, Mark Dunning, J Robin Highley

## Abstract

A key role for inflammation in amyotrophic lateral sclerosis/motor neuron disease (ALS/MND) has been identified. It is vital to assess which central nervous system structures are most affected and which inflammatory processes are responsible in humans. The inflammatory transcriptome was characterized in the cervical spinal cord and motor cortex in post mortem frozen and formalin-fixed paraffin embedded specimens from human sporadic ALS/MND and control cases using the nCounter® Neuroinflammation Panel. Archival data were re-analysed and compared with the nCounter data. Immunohistochemistry was used to examine the inflammatory response in the spinal cord, motor cortex, and regions across the brain and validate changes found at during transcriptomic analyses. In the spinal cord, marked inflammation was observed while less inflammation was detected in the motor cortex. Examination of differentially expressed genes in the spinal cord highlighted *TREM2*, *TYROBP*, *APOE*, and *CD163* as well as phagocytic pathways. In sporadic ALS/MND spinal cord, significant microglial reactivity, and involvement of TREM2, ApoE (encoded by *APOE*) and TYROBP was confirmed, suggesting the involvement of the disease-associated microglial (DAM) phenotype. The corticospinal tracts showed greater inflammation than the ventral horns. The precentral gyrus of ALS/MND again showed less immune reactivity to disease when compared to controls. Finally, in the largest cohort assessed to date, we demonstrate an association between the *APOE* haplotype and ALS/MND risk, age of onset and survival. We confirm associations between *APOE* ε4 and a more aggressive disease; and between ε2 and a less severe disease phenotype. We conclude that while there is widespread inflammation in the CNS in sporadic ALS/MND, this is more marked in the spinal cord, especially the corticospinal tract. The specific markers stress the DAM phenotype as having a key role together with a possible influx of somatic macrophages. In addition, APOE function and genotype may be relevant in ALS/MND.

## Introduction

Motor Neuron Disease (MND) is a fatal neurodegenerative condition, characterised by the progressive degeneration of motor neurons (42) with an incidence of 1.5-2 diagnoses in 100,000 people per year (58). Amyotrophic lateral sclerosis (ALS) is the most common clinical manifestation of MND in adults (accounting for 80-90% of cases) and as such the terms ALS and MND are often used interchangeably. The majority (90-95%) of cases are sporadic. Survival time varies considerably, although 80% of patients survive only two to five years after diagnosis (91). Pathologically, ALS/MND is characterized by motor neuron and pyramidal tract degeneration, together with various proteinaceous inclusions composed principally of TDP-43 (MND-TDP) or cystatin C.

Microglia are the resident immune cells of the central nervous system (CNS), and account for between 5-12% of cells within the brain. Microglial density and function vary greatly with CNS region and age (38, 62). Under physiological conditions, microglia have a ramified morphology with small soma and fine processes (65). Under pathological conditions, the cells react to pathogens and damaged endogenous cells via Damage-Associated Molecular Patterns (DAMPs) with changes both in biological activity (with associated molecular markers) and morphology (30), This results in swollen and shortened cytoplasmic processes (hyper-ramification) and ultimately adopt a large, ‘amoeboid’ morphology. Microglia present multifaceted signalling responses, leading to a complex phenotype. This changes with several factors including the region of the CNS (32), age (29), activating stimuli and disease (28).

The microglial response is critical for CNS protection, through the destruction and removal of damaged or dysfunctional cells or pathogens and the provision of trophic support. This response is highly effective. However, where the initial trigger is not resolved, a destructive cycle of microglial activation and neuron death is initiated: DAMPs that are released by degenerate cells can cause further inflammatory microglial activation resulting in neuronal death (12). This can drive the progression of neurodegeneration (86).

There is good evidence that neuroinflammation is a key feature of human ALS/MND. Firstly, transcriptomic analyses of *post-mortem* spinal cord have highlighted inflammation as one of the most altered pathways (5). At sites of neuronal loss in *post-mortem* tissues, CD68 and Iba1 immunohistochemistry (IHC) reveals microglia to transition from their ramified morphology to an ‘activated’ morphology (16, 17, 37). Microglial activation correlates with the extent of TDP-43 pathology, executive dysfunction, upper motor neuron symptoms, and the rate of disease progression (2, 17). Secondly, *in-vivo* positron emission tomography (PET) shows increased signal in both motor and extra motor regions (89), and an association between microglial activation and cortical thinning and a worse disease phenotype (2). Finally, CSF from ALS/MND patients has high levels of inflammatory cytokines (7, 61, 87), the expression of which has been correlated with the disease progression rate (55, 85, 87).

Human studies have established the presence of a generalised microglial reaction in motor neuron disease/amyotrophic lateral sclerosis. Attempts to specify which of the many molecular behaviours of microglia are relevant have predominantly utilised transgenic animal models of familial (not sporadic) ALS/MND. The most characterised and widely used models are those overexpressing *SOD1* mutations (19, 34, 77, 95). More recently, models have been created which contain transgenes with mutations in other genes (summarised by Lutz, 2018 (59)), as well as in other species including rats (70) and zebrafish (25, 74, 82). Such studies have highlighted both toxic and neuroprotective microglial functions. However, there have been considerable inconsistencies. For example, older studies indicated a tendency towards an early increase in trophic and anti-inflammatory expression prior to symptom onset followed by a switch to a more toxic, proinflammatory phenotype. In contrast, more recent studies have provided evidence indicating microglia express both protective and toxic factors consistently throughout disease (See (5)).

Given the conflicting data from animal studies, it is worthwhile attempting to elucidate specific inflammatory processes using human tissue. There are other good reasons to study MND/ALS-related neuroinflammation in human tissue. Thus, many animal studies have focussed on pathology related to *SOD1* mutations. However, in human MND/ALS, patients with *SOD1* mutations (MND-SOD1), have a different pathological profile, lacking TDP-43 proteinopathy and considerable extramotor disease and have a subtly different clinical profile. It is thus arguably not comparable with ‘classical’ ALS/MND pathology where there is TDP43 proteinopathy (MND-TDP). This issue, in combination with significant interspecies differences in anatomy and immunity (29) raises questions about the validity of animal models to study inflammation in MND. Finally, cultured microglia have also been found to alter their transcriptome and response to stimuli (21), in particular impacting genes associated with neurodegeneration (31).

For this reason, it is crucial to supplement study of microglia in model systems with studies conducted in humans. The study that we report characterises the neuroinflammatory and microglial phenotype in the motor system in human sporadic MND.

We performed a transcriptomic analysis of the motor regions of the CNS (ventral horn of the spinal cord and precentral gyrus – motor cortex) from control and sporadic ALS/MND cases to identify a neuroinflammatory signature. We then proceeded to use immunohistochemistry for microglial and macrophage markers to assess inflammation in the motor system of sporadic MND. Finally, we accessed publicly-available data from the project MinE database (90) and demonstrated very significant relationship between APOE haplotype and MND, that affects both disease risk and severity.

## Methods

### Tissue Collection, preparation, and RNA extraction for NCounter analysis

*Post-mortem* snap-frozen and formalin-fixed paraffin-embedded (FFPE) cervical spinal cord tissue and snap-frozen motor cortex was obtained from the Sheffield Brain and Tissue Bank. Each cohort consisted of 16 sporadic ALS/MND cases of varying survival times from diagnosis to death, and 8 normal control cases (supplementary tables 1-3).

Frozen tissue was brought up to −20°C from −80°C for dissection in a freezer cabinet where he ventral horns and motor cortex were isolated from surrounding tissue by a qualified neuropathologist (JRH). Approximately 20-40mg of tissue was collected per sample. For the formalin-fixed, paraffin-embedded (FFPE) spinal cord tissue, 10 sections of 50µM thickness were cut and the ventral horns isolated as for the frozen spinal cord in a similar manner.

RNA was extracted from frozen tissue using the Zymo Research Direct-zol™ RNA Miniprep kit (Zymo Research, R2050) and from FFPE tissue using the Qiagen® RNeasy FFPE kit (Qiagen, 73504) following manufacturers’ instructions.

### nCounter Gene Expression Assay

The Bruker Spatial Biology (previously NanoString) Sprint Profiler Gene expression assay using the nCounter® Human Neuroinflammation Panel (XT-CSO-HNROI1-12) was used on samples of 100ng of RNA (5µL at 20ng/µL on the advice of nCounter technical staff) according to the manufacturer’s protocols.

The nCounter data were analysed using limma-voom method (80). Briefly, data was normalised using the Trimmed Mean of M Values method, and transformed using the voom function. A linear model for each gene was fitted using the lmFit function. The effect of age and sex were controlled for by adding these as covariates in the analysis. Tests for significance were performed using the eBayes function. Differentially expressed genes were identified with the criteria of Benjamini-Hochberg adjusted p value (false discovery rate)<0.05 and absolute fold change >|1.5|. For the frozen tissue, all cases were kept in analysis. FFPE spinal cord data for three cases (two control and one MND) were discarded due to low count number.

The nCounter Neuroinflammation panel annotates each gene with associated KEGG pathways. The KEGG pathways from all significantly differentially expressed genes were counted to identify those which appeared most frequently. Pathway analysis was not performed on the motor cortex as insufficient genes reached significance following false discovery rate correction.

### Length of Survival Analyses

The association of genes with the length of survival from onset to death in sporadic ALS/MND cases was assessed using Cox proportional hazards regression and the log-rank test in the R survival package (https://cran.r-project.org/web/packages/survival/index.html). Cases were split into high- or low-expressing groups, for each gene, based on the median expression across cases. The effect of age and sex on survival was also analysed using Cox proportional hazards regression. Age and sex did not have a significant influence on survival in this data set.

### Microglial/Macrophage deconvolution

Those genes most associated with microglial/macrophage expression were determined using the online Brain RNASeq tool (https://www.brainrnaseq.org/; (101)). Each of the differentially expressed genes was entered into the database, and the Fragments per Kilobase of transcript per Million mapped reads for the main glial/CNS cell types given. A gene was determined to be microglial/macrophage specific if expression was greatest to the microglial/macrophage population and not highly expressed by any other cell population.

### Comparison of gene expression datasets

The analysis from the frozen spinal cord tissue was compared to the analysis from the FFPE spinal cord tissue. To further assess the robustness of this dataset, we compared our results to the RNAseq data from another laboratory (NCBI sequence read archive SRP064478, (4)). This cohort consisted of six sporadic ALS/MND cases (3 male and 3 female) with a median age of 68.5 years, and eight control cases (4 male, 4 female) with a median age of 66.5 years. One case (ALS4, male) was excluded as this case has a pathogenic *SOD1* mutation.

Raw RNASeq data files were analysed in R. Raw RNAseq reads were quantified and aligned using the Salmon method. The quant files were read and aligned to the human genome. Genes expressed over background in less than two cases were classed as low-expressing and were filtered out. Box plots were used to visualise the spread of the raw data and normalised data, which showed similar distribution across all samples. Heat maps were plotted and PCA performed to look for outliers. However, no clear outliers were identified therefore all cases were kept. Differential expression was performed using the DESeq2 method as described in the R vignette, using the control group as the baseline (57).

### Immunohistochemistry for neuroinflammatory markers in MND/ALS

FFPE *post-mortem* tissue from the precentral gyrus and cervical spinal cord was obtained from the Sheffield Brain and Tissue Bank for immunohistochemistry. Tissue sections were used for assessment of spinal cord (n=28 sporadic ALS/MND, n=14 control; Supplementary Table 4). The motor cortex and underlying white matter were assessed using tissue microarrays (n=63 sporadic ALS/MND, n=7 control; Supplementary Table 5), with two cores sampled from each tissue using the Beecher MTA-1 Tissue Microarrayer (93).

Immunohistochemistry was performed using the avidin-biotin complex (ABC) method on 5µm-thick sections using antibodies, antibody concentrations and antigen retrieval protocols detailed in Supplementary Table 6. Slides were scanned using the Hamamatsu nanoZoomer slide scanner (Hamamatsu, Photonics, Japan). There is some controversy regarding the effectiveness of TREM2 antibodies to appropriately bind the appropriate target (82). In non-CNS tissue, TREM2 is known to be primarily expressed by monocytes (27). To confirm the specificity of the antibody, TREM2 expression was examined in bone marrow and spleen (supplementary figure 1). Both showed expression by small, rounded cells consistent with monocytes.

### Colocalization Serial Staining

APOE expression in the CNS has been well characterised and has been found to synthesised primarily by astrocytes and at lower levels by neurons (94). The cellular morphology observed here was consistent with this. As TREM2 and TYROBP are less well characterised, we performed double staining to determine which cell type was expressing these markers. Slides were dewaxed, rehydrated through alcohols, and endogenous peroxidases were blocked in 3% hydrogen peroxide for 20 mins. The slides underwent antigen retrieval, and a standard ABC immunohistochemistry protocol. The first protein of interest was visualised using the ImmPACT AMEC red substrate kit (Vector Laboratories, SK-4285). Slides were washed in tap water and mounted using aqueous mounting media, dried and digitised. Following this, slides were de-coverslipped and the dye was dissolved in graduated alcohols. Unbound avidin and biotin from the first stain was blocked using avidin and biotin blocking solution (Vector Laboratories, SP-2001). The slides were then rehydrated and stained using a standard ABC protocol using a second primary antibody, visualised with DAB, coverslipped and rescanned.

### Image Analysis

Visiopharm (Hoersholm, Denmark) image analysis software was used to quantify immunoreactivity using a previously-developed APP which assesses area density (percentage of a region of interest that is positive for immunoreactivity) (96). Following pre-processing, APPs were trained specifically for each protein of interest, using the Visiopharm training wizard.

In spinal cord sections stained for TREM2, motor neurons showed evidence of lipofuscin staining by DAB which the Visiopharm software was unable to differentiate from true signal. To prevent bias in quantification – as sporadic ALS/MND cases will have fewer motor neurons compared to controls, neurons were excluded from analysis using a filter which re-classify DAB positive areas larger than 200µM as areas in background.

In the spinal cord, specific regions were selected for quantification: the ventral horns, the lateral corticospinal tracts, and the dorsal column. The latter is an ascending sensory tract that we expected to be less affected by MND pathology. All statistical analysis was performed using GraphPad Prism (versions 7,8,9). For all statistical analysis the significance value was set at alpha<0.05.

As many data sets were not normally distributed (by Shapiro Wilk), non-parametric statistics were used for intergroup comparisons: The Kruskal-Wallis test followed by post hoc Mann-Whitney *U* tests were used for most intergroup comparisons.

To examine the relationship with patient survival, sporadic ALS/MND cases were split into fast or slow progressing groups based on the median length of survival for each cohort again using Kruskal-Wallis followed by post hoc Mann-Whitney U tests.

### Genotyping of APOE

Phased genotyping data for rs429358 and rs7412 was obtained from the latest ALS/MND GWAS consisting of 29,612 ALS/MND patients and 122,656 controls (90). Samples were assigned to either heterozygous or homozygous for the APOE haplotypes ε4, ε3 and ε2. Analysis of the relationship between ALS/MND status and APOE genotype was conducted by multivariable logistic regression using sex and the first 10 principal components of genetic variation as covariates. For this analysis, homozygous and heterozygote individuals were considered equivalent. Cox regression was used to assess the relationship with survival and age of disease onset using sex, age (for survival only), and the first 10 principal components of genetic variation as covariates.

## Results

### The transcriptomic profile shows considerable inflammation of the spinal cord in MND/ALS and highlights the ApoE-TYROBP-TREM2 pathway

The nCounter Neuroinflammation Panel was used to assess the spinal cord ventral horn in two tissue cohorts (one snap-frozen and one FFPE). Significantly more genes were upregulated (76, frozen tissue dataset and 62 FFPE tissue dataset) than down-regulated (13 frozen tissue and 38 FFPE tissue; Figure 1, Supplementary Tables 7-10).

**Figure 1:**
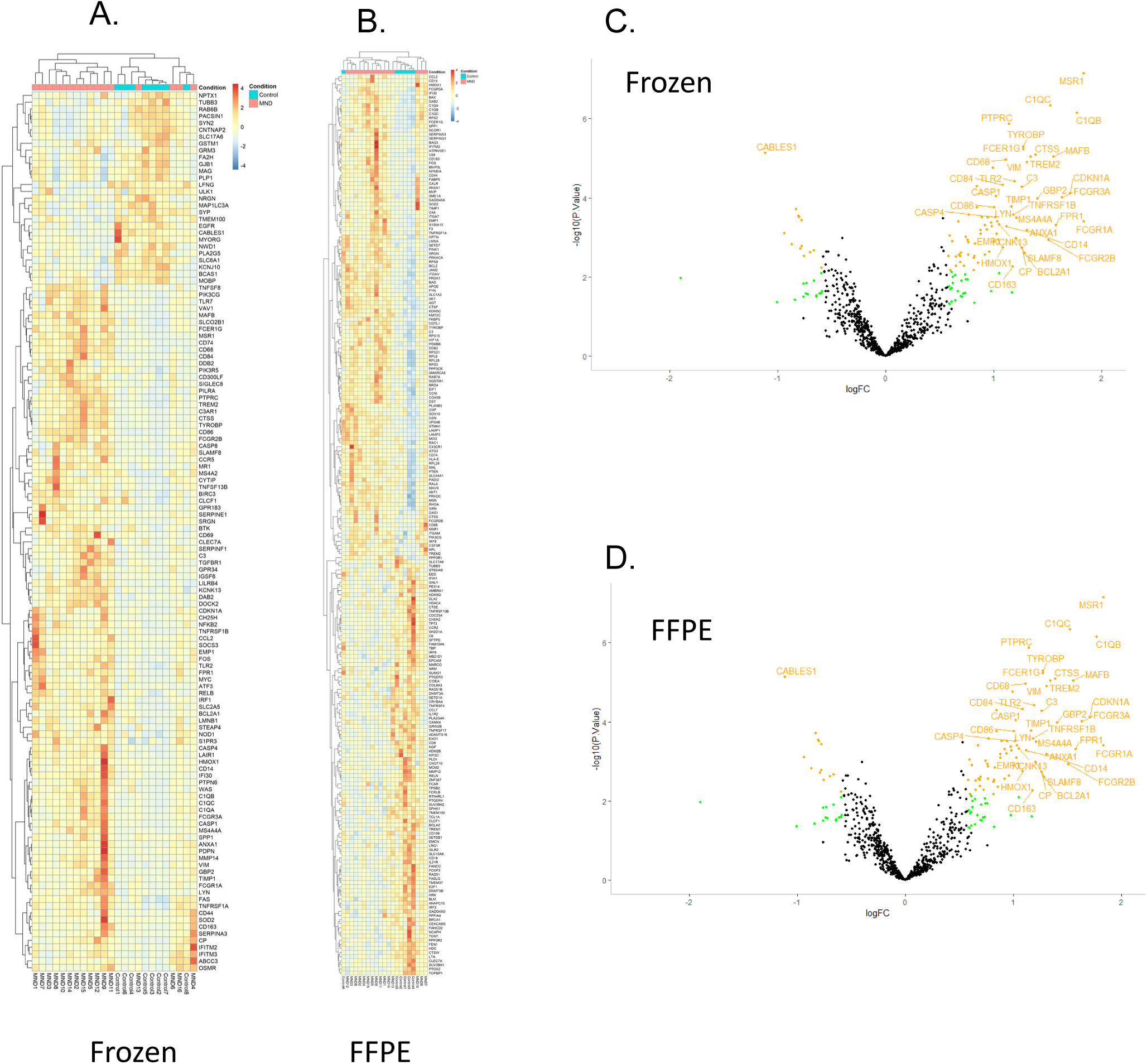
sMND is associated with an Increase in Inflammatory Signalling in the Spinal Cord – Frozen and FFPE Tissue. A. Heatmap displaying the normalized and row-scaled expression of 128 differentially expressed genes (p < 0.05 and FC > 1.5) between MND and neurologically healthy cases. Hierarchal clustering grouped the majority of MND cases and control cases, shown on the x axis. Genes are shown on the y axis. B. Heatmap displaying the normalized and row-scaled expression of 219 differentially expressed genes (p < 0.5 and FC > 1.5) between MND and neurologically healthy cases. Hierarchal clustering grouped the majority of MND cases and control cases, shown on the x axis. Genes are shown on the y axis. C. Volcano plot showing fold change against the significance value. Of all the significant genes, 72 were upregulated, and 13 genes were down regulated (Green Points p<0.05; FC>1.5; Orange Points FDR adjusted p<0.05; FC>1.5, labelled points FDR adjusted p<0.05; FC>2). D. Vo_1_l_5_cano plot showing fold change against the significance value. Of all the significant genes, 72 were upregulated, and 13 genes were down regulated (Green Points p<0.05; FC>1.5; Orange Points FDR adjusted p<0.05; FC>1.5, labelled points FDR adjusted p<0.05; FC>2). Inflammation in the motor cortex was studied using the nCounter platform in one snap frozen tissue cohort (n=16 sporadic ALS/MND cases and 8 neurologically healthy controls for each dataset). In contrast to the spinal cord, none of the 770 genes assessed were significantly altered following false discovery rate correction (Figure 2).

The fold changes of all 770 genes (including those that were not significantly affected by disease) from frozen spinal cord data set was significantly correlated with the fold changes from both the FFPE dataset (R=0.148, p<0.001) and another RNASeq spinal cord MND dataset from the literature used for validation (18) (R= 0.25, p<0.0011). Importantly, all three datasets (our frozen and FFPE nCounter datasets, and the RNAseq dataset (18)) showed upregulation of *APOE*, *TYROBP* and *TREM2*. These three components form a pathway with a key role in inflammatory regulation and can induce a disease-associated state in microglia (see below).

Of the 89 differentially-expressed genes from the frozen dataset that were known to have cell-specific expression (101), 50 genes were known to be primarily expressed by microglia or macrophages. The KEGG pathways associated with the differentially expressed transcripts from sporadic ALS/MND spinal cord were ranked by the number of genes associated with each pathway (see Supplementary Table 11).

By Cox Proportional Hazards Regression, 23 and 17 genes were associated with longer survival in the frozen and FFPE datasets respectively. Shorter survival was associated with 21 and 23 genes in the frozen and FFPE datasets respectively (Supplementary tables 12-15). Functionally, genes associated with longer survival were involved in both the adaptive and innate immune response, cytokine signalling, growth factor signalling, autophagy, and apoptosis. Genes associated with shorter survival had overlapping functions namely innate and adaptive immunity, cytokine signalling, growth factors and microglial function.

In summary, RNA-based gene expression analysis shows concordance between nCounter datasets obtained from FFPE and frozen tissue, and the nCounter analyses are consistent with an archival RNAseq dataset from the literature. The inflammation in the spinal cord appears more severe than in the motor cortex in MND. The analysis highlighted several key inflammatory processes including APOE-TYROBP/DAP12-TREM2 signalling.

### Immunohistochemistry for inflammatory markers in sporadic ALS/MND

Immunohistochemistry was performed to elucidate and confirm the findings from the nCounter-based investigation above: Ionized calcium binding adaptor molecule 1 (IBA1, a fairly universal marker of microglia and monocytes (13)), MHCII/ HLA-DR (a marker of activated microglia (13), and CD68 (a marker of microglial phagocytosis (43)) were assessed. In addition, Cluster of Differentiation 163 (CD163, primarily expressed by perivascular macrophages (14)), was assessed as this highlighted by the nCounter data.

We tested the following hypotheses:

1. Expression of microglial and macrophage markers (IBA1, CD68, HLA-DR and CD163) will be increased in sporadic ALS/MND compared to controls. Microglia will display activated morphology characterised by swelling of the processes and cell body, and/or the presence of amoeboid microglia in sporadic ALS/MND CNS regions. This will be more marked in spinal cord than motor cortex. Within the spinal cord, there will be greater inflammation in motor regions (ventral horns and corticospinal tracts) than sensory regions.
2. ApoE, TREM2 and TYROBP expression will be increased in the spinal cord in sporadic ALS/MND cases compared to control cases.
3. There will be greater a microglial response and increased expression of target proteins in the spinal cord overall compared to the motor cortex brain in sporadic ALS/MND cases.
4. Expression of proteins of interest will be associated with survival, particularly in the spinal cord.

### There is substantial spinal cord inflammation in sporadic ALS/MND, that is variable in anatomical extent and severity

In qualitative terms, IBA1, CD68 and HLA-DR labelled microglia and perivascular macrophages in both control and sporadic MND/ALS cases. However, in sporadic ALS/MND, IBA1^+^ microglia had a more reactive/activated morphology characterised by thicker processes and larger soma, with some cells acquiring an amoeboid form. This was most marked in the motor regions (ventral horns and corticospinal tracts; Figure 3 and Supplementary Figures 2-4). This reaction showed variation in anatomical extent, severity of inflammation and cellular morphology within the MND cohort. While the inflammation was most prominent in motor regions, for some cases, the reaction was present throughout most of the spinal cord, albeit with lesser involvement of the dorsal columns (Figure 4). Thus, classically sensory tracts such as the spinothalamic tract were involved in some individuals. HLA-DR showed more florid inflammation in terms of the degree of labelling compared to other markers (Figure 5c and Supplementary Figure 4).

**Figure 2:**
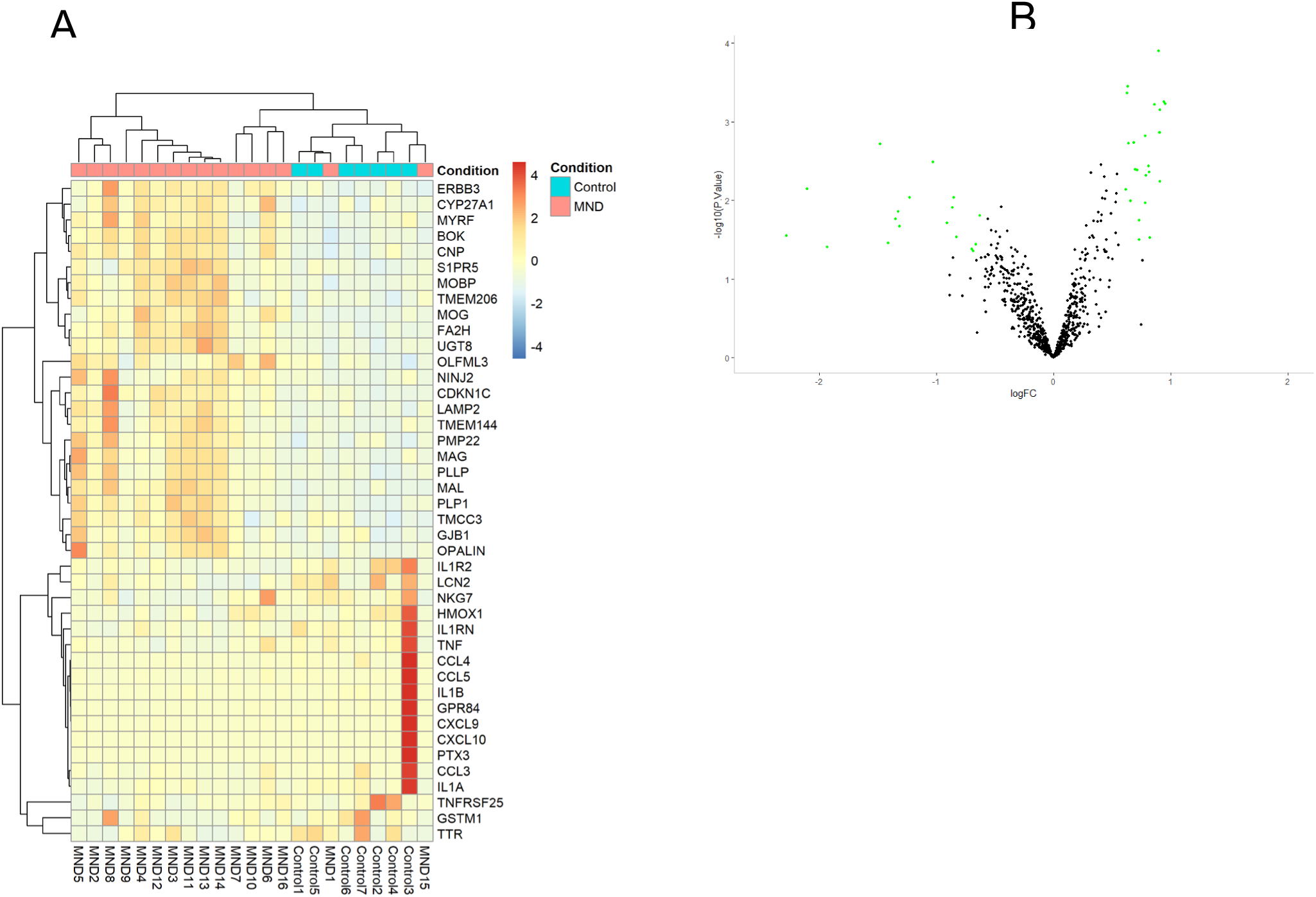
The Motor Cortex Shows Little Inflammatory Signalling in sMND. A. Heatmap displaying the normalized and row-scaled expression of 42 differentially expressed genes (p < 0.05 and FC > 1.5) between MND and neurologically healthy cases. MND and control cases show good separation with some overlap of cases. B. Volcano plot showing fold change against the significance value. Green Points p<0.05; FC>1.5. No genes reached significance following FDR correction.

**Figure 3.**
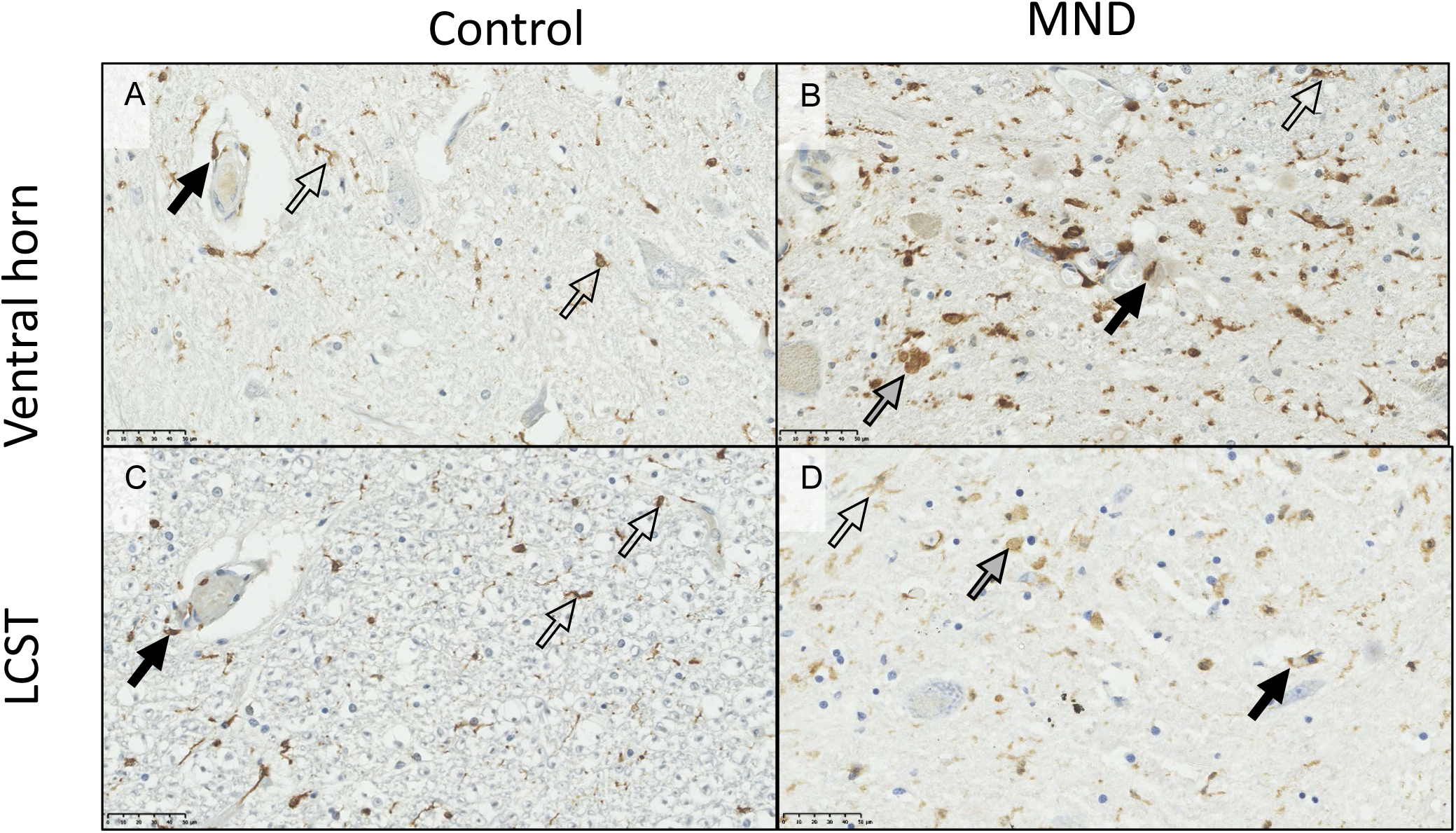
IBA1 In the spinal cord of controls and ALS/MND cases. IBA1 labels perivascular macrophages (black arrows), and microglia (open arrows). In control spinal cord, microglia tend to be ramified. In MND spinal cord, perivascular macrophages were swollen in some cases compared to control. Microglia tended to show a mixture of ramified (open arrows) and amoeboid (grey arrows) morphology. A, control ventral horn; B MND/ALS ventral horn; C, control lateral corticospinal tract (LCST); D, sMND/’ALS corticospinal tract. Scale bars = 50μm

**Figure 4:**
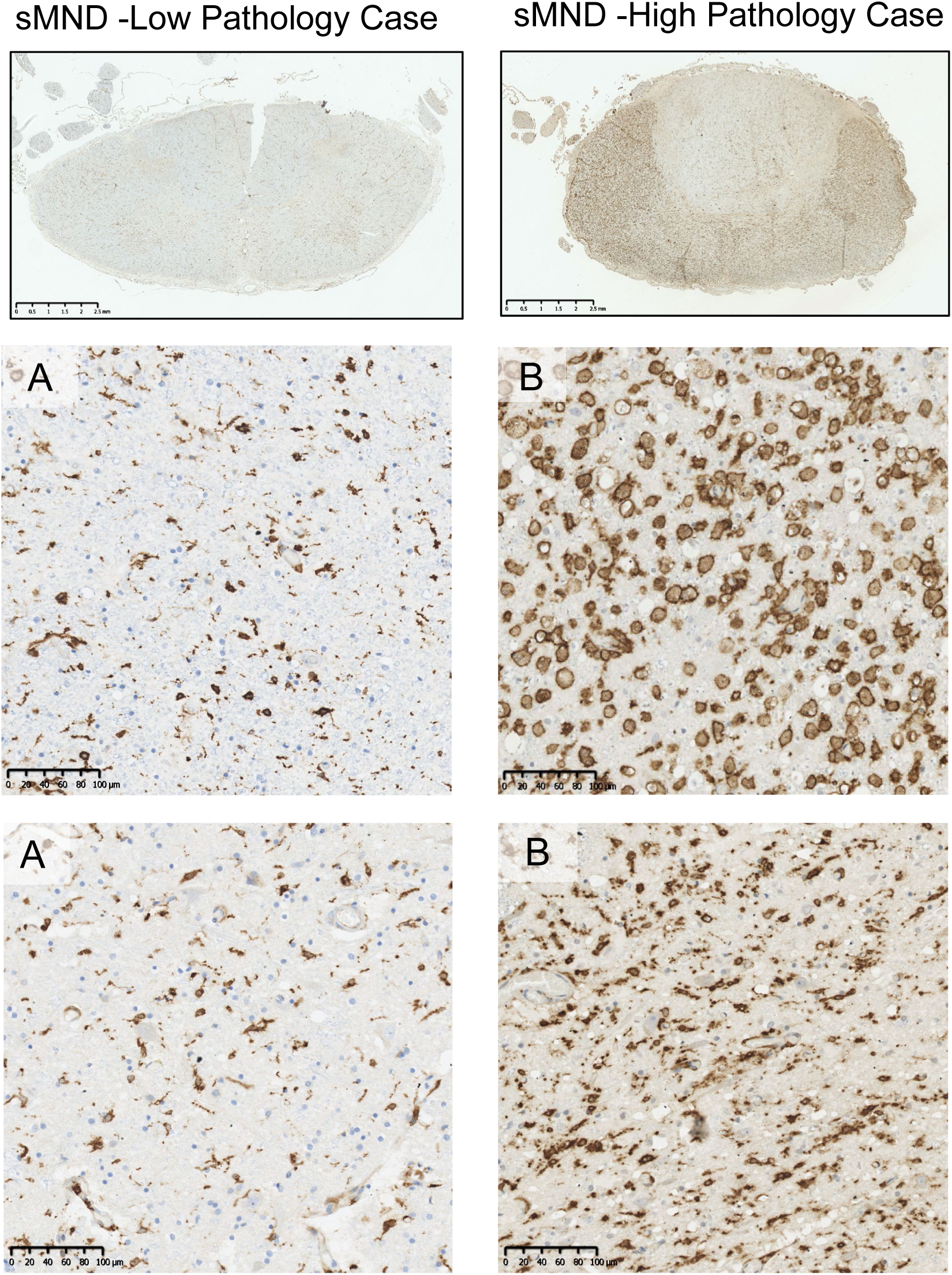
Heterogeneity of HLA-DR Immunoreactivity in sMND Cases. In the spinal cord, sMND cases varied greatly in the level of HLA-DR staining present. Both cases (A & B) are sMND cases. However, as visible from the low magnification image, these cases showed very different levels of HLA-DR positive microgliosis. A1 and B1 show higher magnification images of the lateral corticospinal tract for these cases. In the low pathology case (A1) microglia were rounded (when compared to control) and showed thickened processes. In the high pathology case (B2) microglia were completely rounded and showed much denser staining. A2 and B2 show higher magnification images of the ventral horn. In case A (low pathology) microglia again showed thick processes, swollen cell bodies and an increased number of HLA-DR positive microglia. In case B, HLA-DR positive microglia were much rounder, many becoming ameboid, and microgliosis was much more severe. A and B scale bars =2.5mm; A1, A2, B1 and B2 scale bars =100µm.

**Figure 5:**
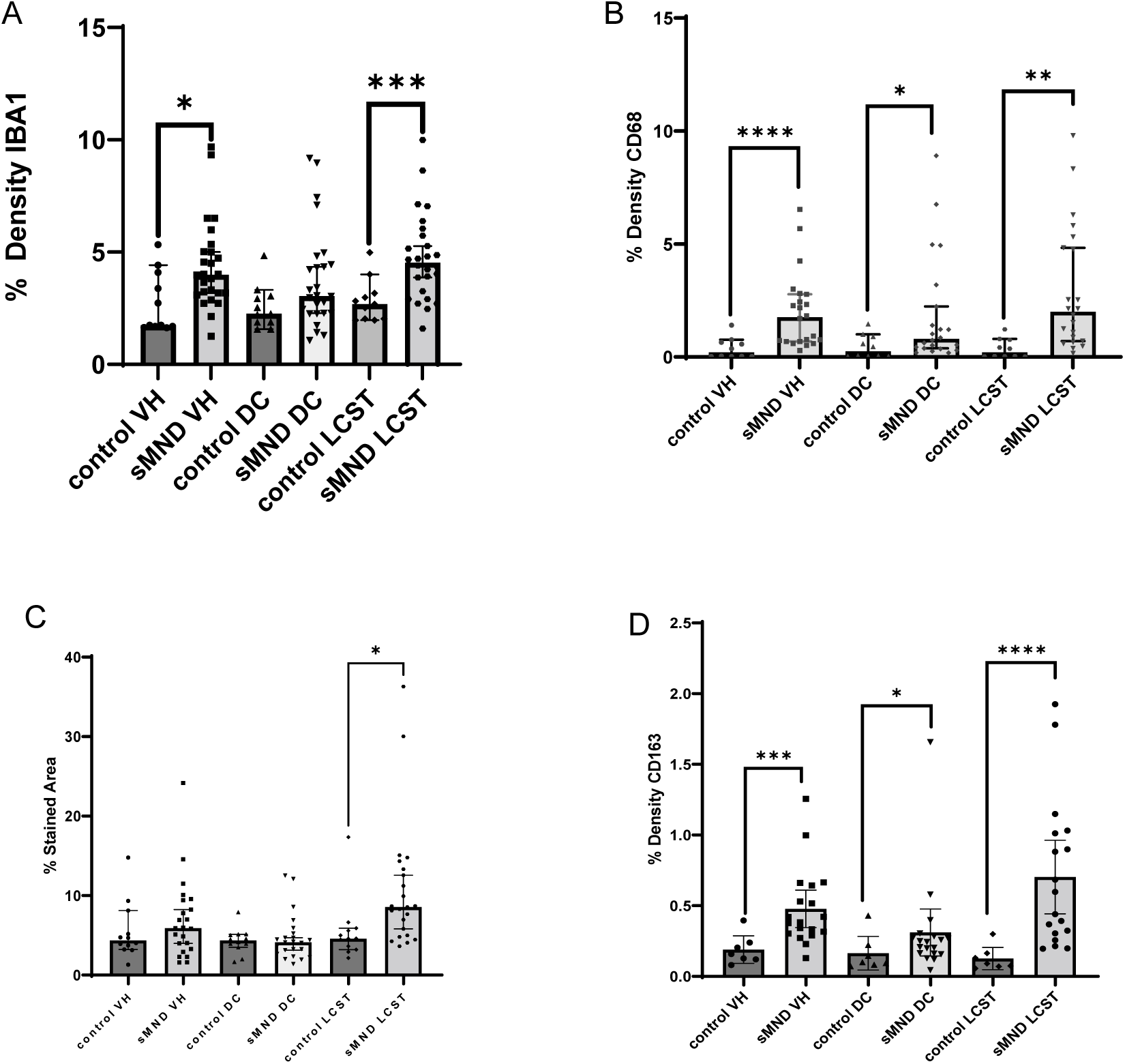
Area density for a variety of monocyte/microglial markers in MND/ALS spinal cord. There is greater inflammation in the motor structures of the cord, namely the ventral horns (VH) and lateral corticospinal tracts (LCST) in MND/ALS as shown by IBA1 (A), CD68 (B), HLA-DR (C) and CD163 (D). There is some lower level inflammation seen in the sensory dorsal columns when assessed by CD163 and CD68.

CD163 was largely confined to perivascular macrophages with minimal microglial expression in control spinal cord (Supplementary Figure 5). In MND/ALS cases, there was a variable increase in the number of positive cells in the ventral horns and corticospinal tracts both by perivascular macrophages, and also by small numbers of cells within the spinal cord parenchyma.

By image analysis, the ventral horn, dorsal column, and lateral corticospinal tract showed significant upregulation of expression of IBA1, CD68, HLA-DR and CD163 in the lateral corticospinal tracts in MND/ALS compared to controls. Significant upregulation was seen in the ventral horn for Iba1, CD68 and CD163, but not HLA-DR. There was also significant upregulation in the dorsal column for CD68 and CD163 (Figure 5). MND/ALS patients with shorter survival had greater HLA-DR expression in the ventral horns and lateral corticospinal tracts (*P*=0.0148 and 0.005 respectively).

### The Apolipoprotein E-TREM2-TYROBP pathway is up regulated in motor components of the spinal cord

ApoE immunohistochemistry revealed expression throughout the spinal cord that was more marked in grey than white matter. While there was labelling of the cord parenchyma, there was especially strong labelling in glial cells (Figure 6, supplementary figure 6). A subset of motor neurons was also strongly stained. In MND/ALS cases, there was a greater number of glial cells expressing ApoE in the corticospinal tract and in the ventral horns. The proportion of motor neurons that were positive or negative for ApoE did not differ between MND/ALS cases and controls.

**Figure 6:**
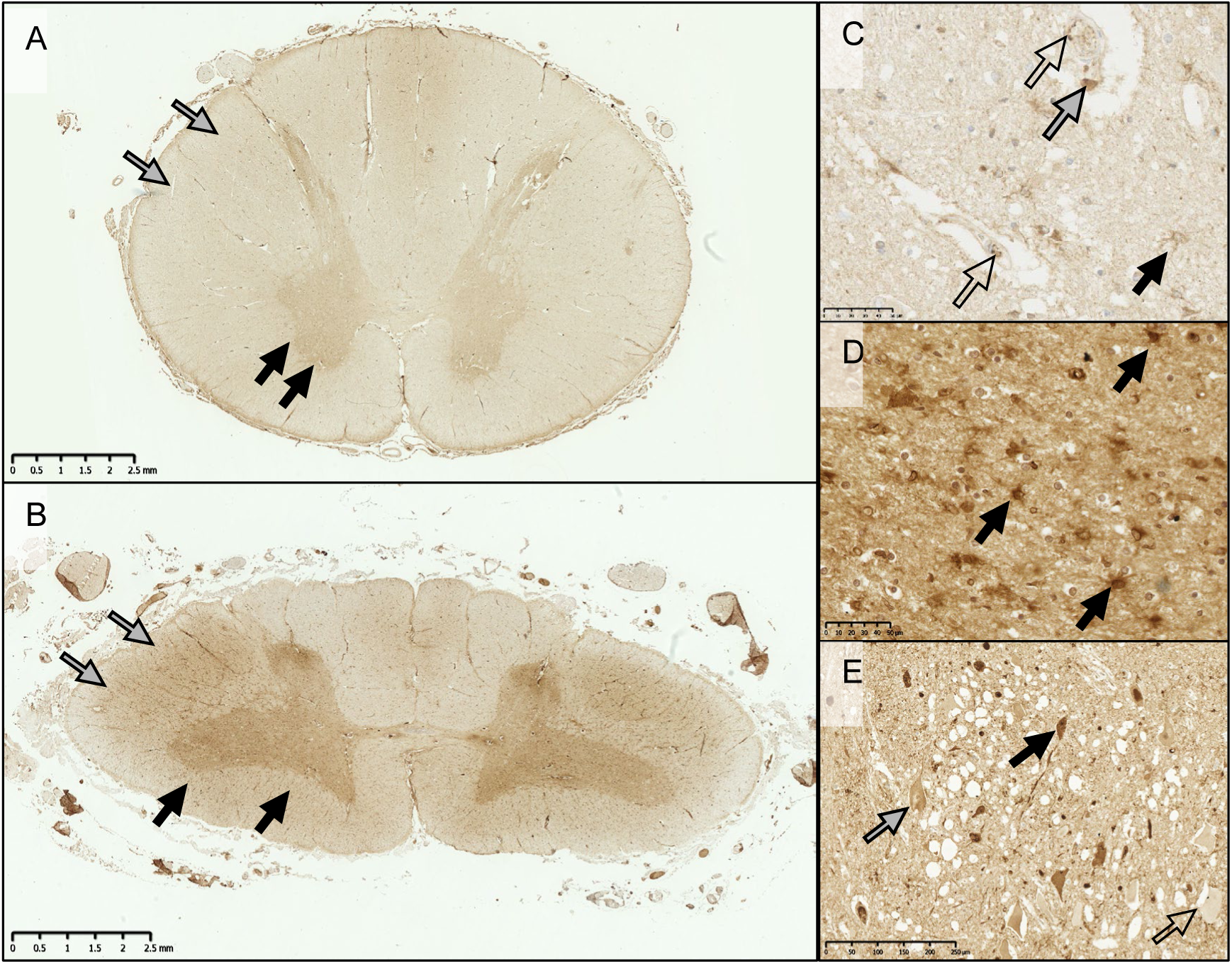
APOE Immunoreactivity in the Spinal Cord. In control (**A**) and MND/ALS (**B**) spinal cord, at low power ApoE is seen in higher levels in the grey matter compared to the white matter. In MND/ALS cases, ApoE is increased in the corticospinal tract (grey arrows), and ventral horns (black arrows). At higher power (C, control ventral horn; B, MND/ALS ventral horn), ApoE is present in endothelial cells (open arrows), perivascular macrophages (grey arrows) and parenchymal glial cells (black arrows) that have the appearance of astrocytes, as well as the background neuropil. There is also variable motor neuron staining: some having high signal (black arrow), others had similar signal to the surrounding parenchyma (grey arrows) and a small number of neurons had no immunoreactivity (open arrows). This differential ApoE expression varied greatly between cases and was not associated with disease/control status. Scale bars: A,B=2.5 mm; B,C=50 μm; D=100 μm.

Digital image analyses showed significantly greater cellular ApoE area density in the lateral corticospinal tract (U=20, p=0.0003) and ventral horns (U=32, p=0.0036) but not the dorsal columns in MND/ALS (U=51, p=0.0949). TYROBP (also known as DAP12), in common with ApoE, was present in neuropil, neurons, glia and blood vessels with greater expression in grey than white matter in both control and sporadic ALS/MND groups (figure 7, supplementary figure 7).

**Figure 7:**
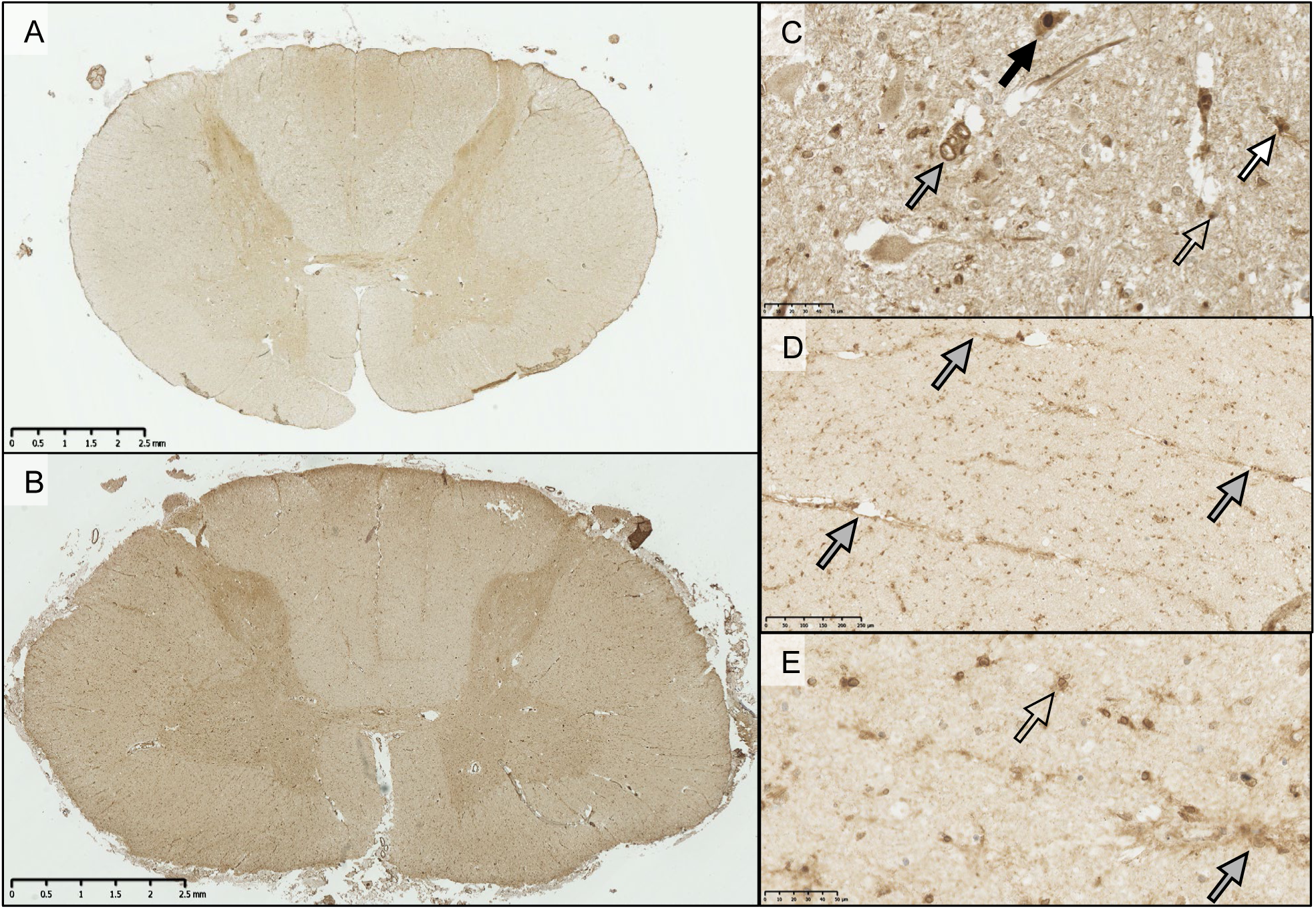
Expression of TYROBP in the Spinal Cord. In control (**A**) and MND/ALS (**B**) spinal cord at low magnification, TYROBP immunoreactivity was present in the neuropil, in higher levels in the grey matter compared to the white matter. Glial staining in the white matter, particularly in the corticospinal tracts was increased in MND/ALS cases. At higher magnification (**C**), TYROBP labelled motor neurons in the ventral horn (black arrow), and blood vessels (grey arrow), as well as various ramified (white arrow) and unramified glia (open arrow). Image taken from the ventral horn. In MND/ALS ventral horn (D,E), there was greater TYROBP expression in perivascular cells. Scale bars: A,B=2.5 mm; D=250μm; C,E=50μm

In sporadic ALS/MND cases, microscopy revealed an increase of both perivascular macrophage and interstitial glial TYROBP staining, especially in the corticospinal tracts and the ventral horns. Ramified cells tended to be observed in the ventral horns and more amoeboid cells in the corticospinal tracts (supplementary figure 7). Glial upregulation was especially marked in perivascular locations (figure 7). Image analysis confirmed increased TYROBP in the ventral horns (U=11, p=0.009) and corticospinal tracts (U=12, p=0.018), but not dorsal columns in MND/ALS (U=20, p=0.075; figure 9). There was no relationship with survival time (p=0.932).

TREM2 expression was seen in a few perivascular macrophages, a variable proportion of motor neurons and a few glial cells in the parenchyma (figure 8, supplementary figure 8).

**Figure 8:**
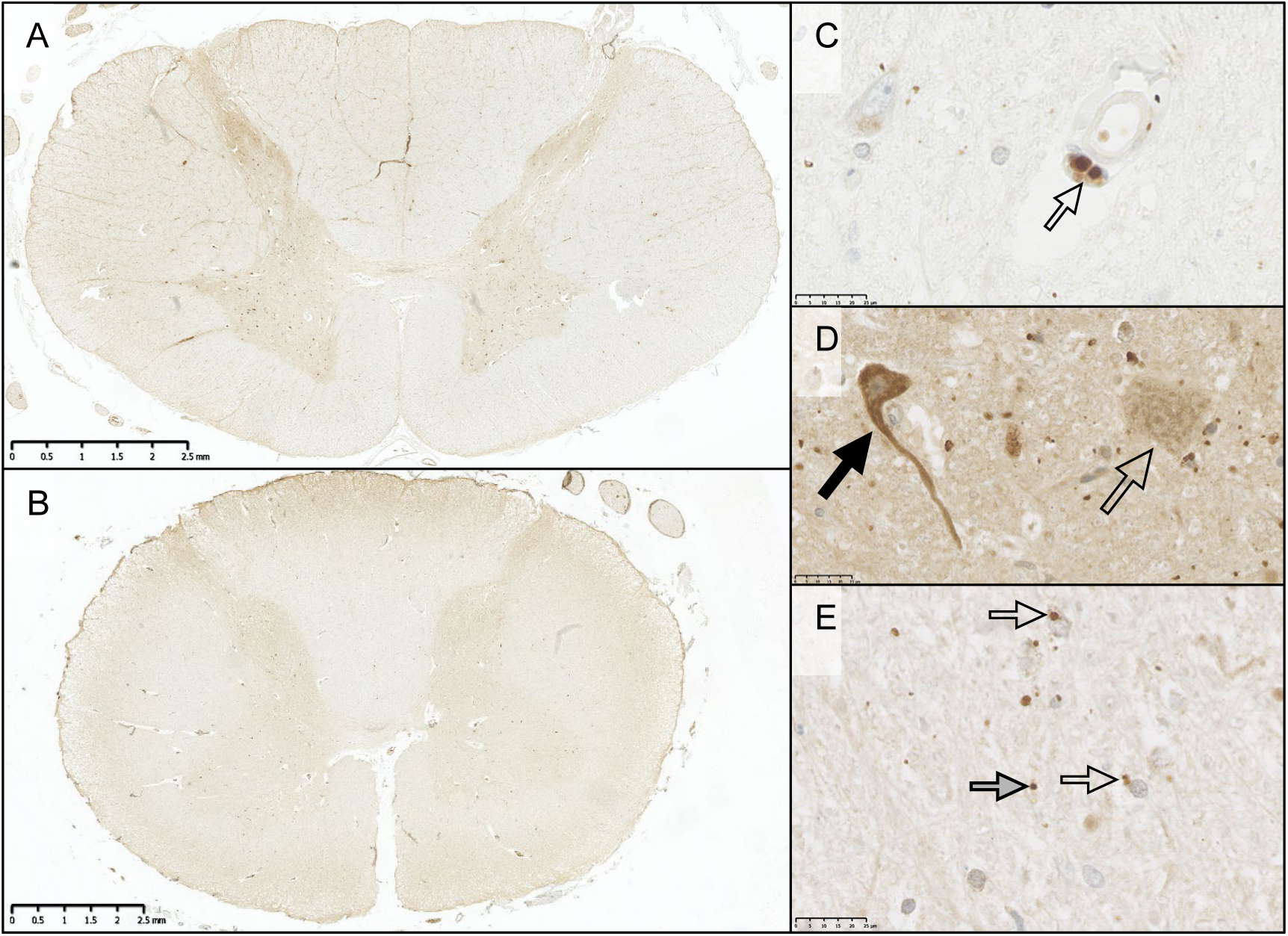
TREM2 Expression was Minimal in the Parenchyma. TREM2 immunoreactivity was minimal in the parenchyma, although some cases did show slightly greater labelling of the grey matter compared to white. At low power, there was little difference between control cases (A) and sMND cases (B). At higher power, TREM2 was seen in a few small, rounded cells likely perivascular macrophages (C). The majority of neurons showed minimal TREM2 signal (open arrow, D). However, TREM2 immunoreactivity did label a small number of motor neurons more strongly in the ventral horn (black arrow). These were not present in all cases, and did not appear to be associated with either sMND or control cases specifically. In the parenchyma, small TREM2+ve granules were observed. Some granules were associated with glial or monocyte cell (open arrow). Others were not (grey arrow). Scale bars: B=2.5mm; C,D,E = 25μm.

**Figure 9:**
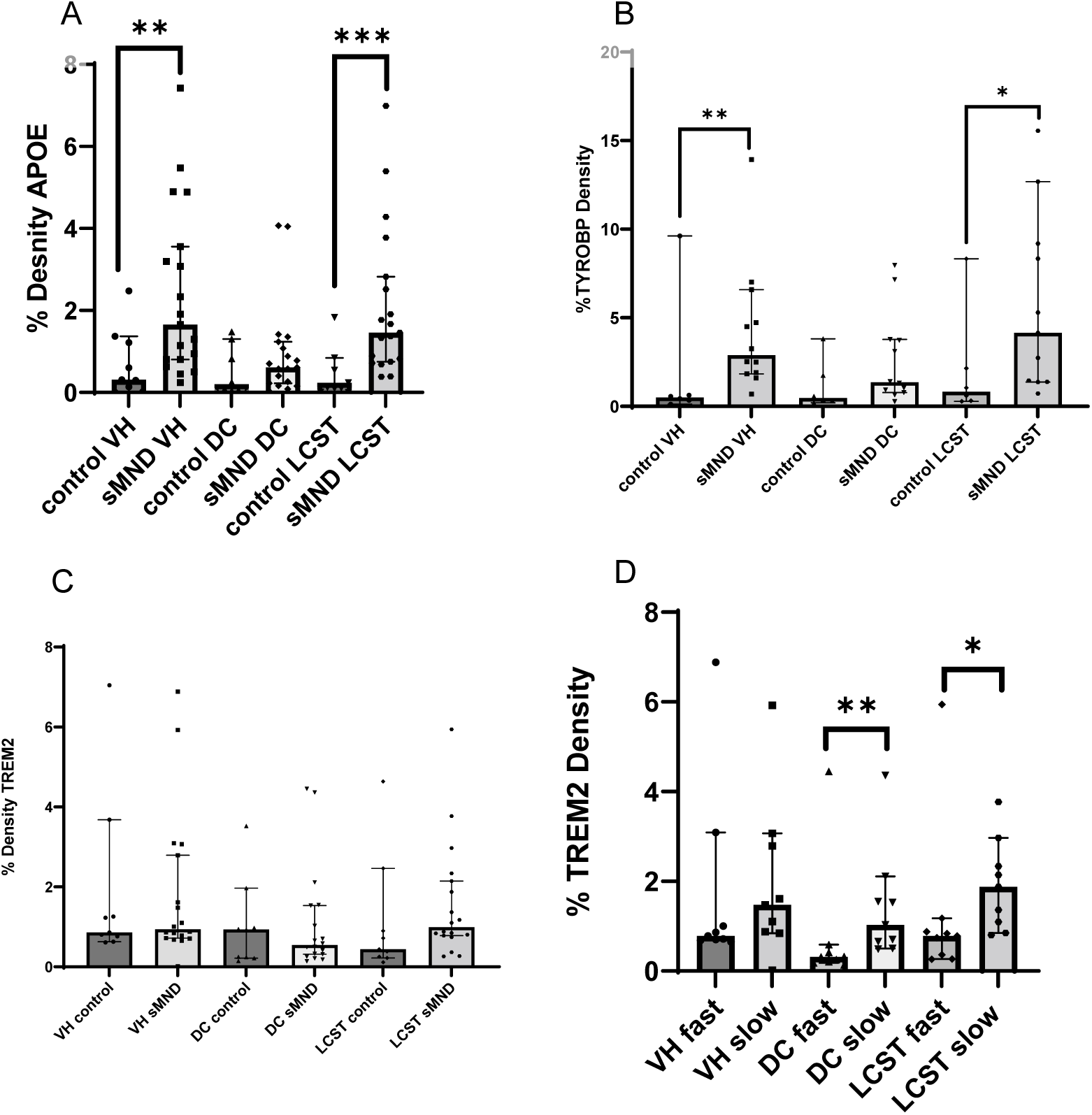
Area density for a variety of ApoE, TRYOBP and TREM2 in MND/ALS spinal cord. There is greater expression of ApoE (A) and TYROBP (B) in the motor structures of the cord, namely the ventral horns (VH) and lateral corticospnal tracts (LCST) in MND/ALS. There were no such intergroup differences in TREM2 (B). However, excess TREM2 expression was associated with slower disease progression (D).

There was greater expression in the grey compared to white matter. Control and MND/ALS cases showed similar patterns of TREM2 expression). Image analysis found no difference between groups in the degree of TREM2 expression (H(5)=8.449, p=0.133; figure 10).

**Figure 10:**
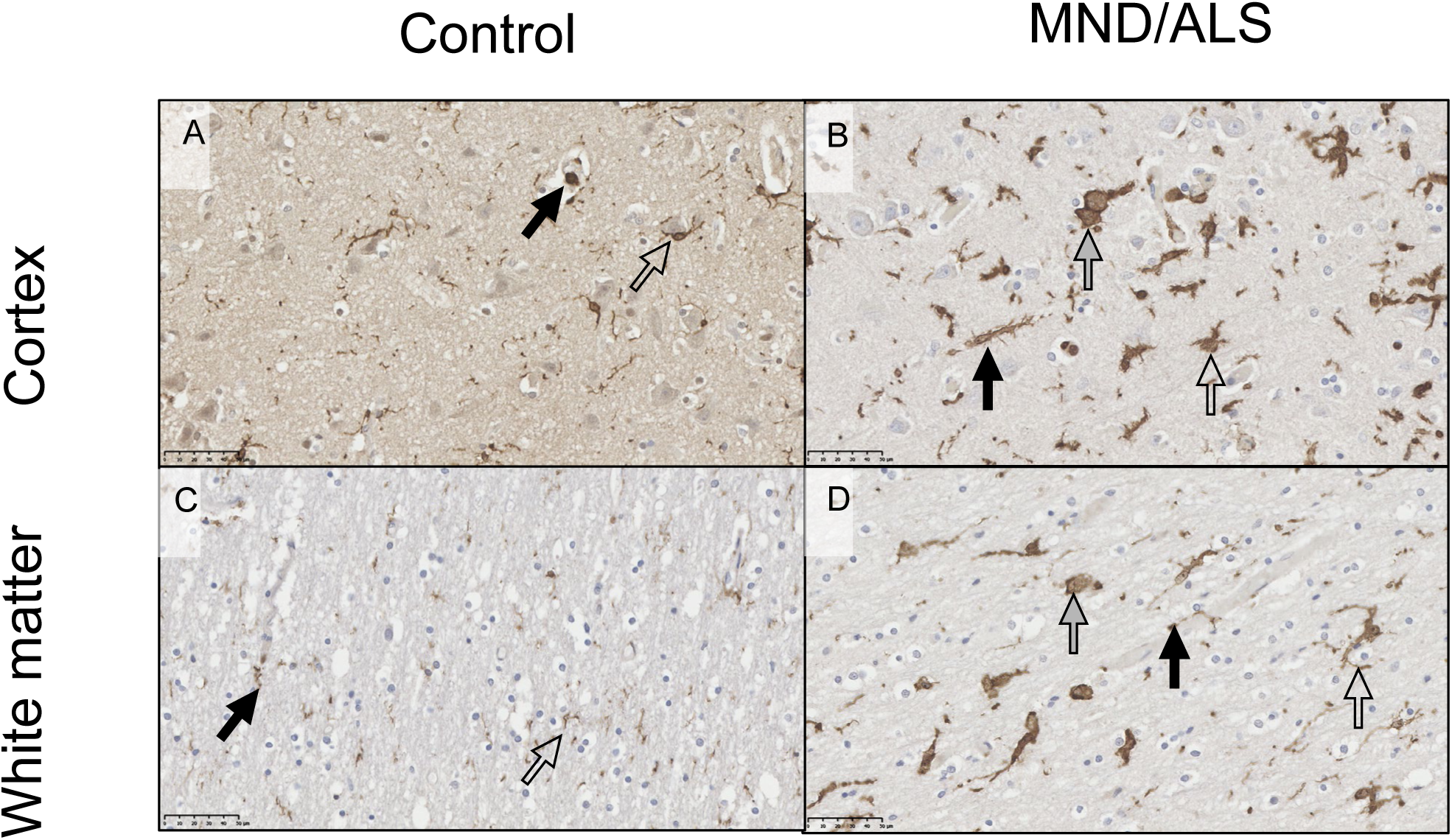
IBA1 in Motor Cortex. IBA1 labelled perivascular macrophages (black arrows) and microglia (open arrows). In control cases, microglia were ramified with small cell bodies and fine processes. Similar patterns of expression were observed in the grey matter (**A**) and white matter (**C**). In MND/ALS (**B,** cortex; **D,** white matter), microglia had an activated morphology, with thicker cell bodies and swollen processes (open arrows), some microglia had also transitioned to the fully amoeboid state (grey arrows). Perivascular macrophages are labelled with black arrows. Expression was similar in grey matter (C) and white matter (D). Scale bar = 50μm.

There was a significant relationship between longer survival and greater expression in the white matter tracts (dorsal column U=11, p=0.008; corticospinal tract, U=15, P=0.024), but not the ventral horn (U=27, p=0.258) by digital image analysis.

To determine which cells were expressing ApoE, TYROBP and TREM2, sections of spinal cord from MND/ALS individuals were serially immunostained for ApoE, TYROBP or TREM2 then IBA1 (supplementary figure 9). This revealed that most of the cells that were strongly positive for these markers were IBA1-positive microglia and perivascular macrophages.

### The neuroinflammatory response in the motor cortex is less than the spinal cord in MND

In both sporadic ALS/MND and control cases, as was observed in the spinal cord, IBA1, CD68 and HLA-DR label microglia and perivascular macrophages with considerable individual variation in the number of cells labelled. CD163 was largely confined to perivascular macrophages (supplementary figure 10).

In control cases, microglia were ramified with fine processes in both white and grey matter. In sporadic ALS/MND cases, there were activated microglia with thickened processes as well as some amoeboid cells in both the white and grey matter (Figure 10). In contrast to the spinal cord, CD163 was limited to perivascular macrophages with no labelling of parenchymal cells (supplementary figure 10). While there were morphological differences in microglia, there were no intergroup differences in the area density of IBA1, HLA-DR or CD163 (p≥0.141). There was no relationship between of IBA1 or HLA-DR area density and survival (H[3]=1.015, p=0.798). While Kruskal Wallis suggested relationships between CD68 and diagnosis (p=0.0315) and survival in MND cases (p=0.0006), this was not supported by post hoc tests (all p≥0.319).

ApoE (supplementary figure 11) and TYROBP (supplementary figure 12) immunohistochemistry in the motor cortex showed a similar pattern to that of the spinal cord: In controls, there was variable expression in neurons glia and vascular structures with significant expression of ApoE in the neuropil that was less marked for TYROBP. For TYROBP, the area density in both cortex and white matter was significantly greater in sporadic ALS/MND than control cases in both grey and white matter.

There was an apparent relationship between TYROBP staining density and patient survival by Kruskal-Wallis (H(3)=12.44, p=0.006). However, post hoc Mann-Whitney U tests showed no effect of survival in either grey or white matter (p≥0.077).

In the motor cortex, TREM2 expression was largely confined to perivascular macrophages with minimal staining of neurons and negligible effect of disease (supplementary figure 13). There were no differences between control and MND/ALS cases.

As noted above, the transcriptomic analyses of mRNA by nCounter revealed the spinal cord to be an area of much greater inflammatory signalling than the motor cortex. We wished to investigate if this was borne out at the protein level by examining our immunohistochemistry area density data by comparing the white matter of the precentral gyrus with the lateral corticospinal tracts and the motor cortex with the spinal cord ventral horn. This revealed significantly greater expression density in the spinal cord than the motor cortex for Iba1 (U=7, p=0.006), HLA-DR (U=0, p=0.0001), TYROBP (U=0, p=0.002) and TREM2 (U=0, p=0.0004) for control white matter, HA-DR (U=0, p=0.0001), CD163 (U=4, p=0.014) and TREM2 (U=0, p=0.0004) for control grey matter and all markers in the spinal cord compared to precentral gyrus in the ALS/MND cases (all p≤0.012).

### The APOE ε3 haplotype is over-represented within sporadic ALS/MND patients, while ε2 and ε4 are under-represented

Having established upregulation in components of the ApoE-TYROBP/DAP12-TREM2 pathway in MND and an association between TREM2 and disease severity (as indexed by survival time), we were interested in the genotype of *APOE*. This was assessed using data from project MinE, including 29,612 ALS patients and 122,656 controls (77).

ALS patients were significantly enriched with the APOE ε3 haplotype (OR=1.1, beta=+0.1, p=0.02) but depleted of both the ε2 haplotype (OR=0.82, beta=-0.2, p=2.9e-11) and ε4 haplotype (OR=0.93, beta=-0.07, p=3.8e-5).

In a subset of ALS patients for which survival data was available (n=4,897), the ε4 haplotype was associated with shorter survival (coef=0.08, p=0.04) but not age of onset (p=0.7). Over the disease course, carrying the ε4 haplotype compared to not carrying the haplotype was associated with a hazard ratio (HR) of 1.08 indicating an increased risk of death. The ε2 haplotype was associated with a significantly later age of onset (coef=-0.096, p=0.02), but not length of survival. Carrying the ε2 haplotype compared to not carrying the haplotype was associated with a hazard ratio of 0.91 indicating a reduced risk of disease onset. The ε3 haplotype was associated with neither age of onset nor survival.

## Discussion

The inflammatory mRNA transcriptomic profile of ALS/MND was characterised in the *post-mortem* spinal cord and motor cortex. Two datasets were generated for the spinal cord using the nCounter platform. These were compared with a third RNAseq dataset from the literature. There was good correlation between the three datasets with the inflammatory response in the spinal cord greater than that in the motor cortex. A number of neuroinflammatory pathways were highlighted in the spinal cord, notably all three elements of the ApoE-TYROBP-TREM2 pathway.

Following this, immunohistochemistry was used to elucidate neuroinflammation at the protein level. This confirmed that inflammation is far greater in the spinal cord than in the motor cortex and showed marked inter-individual variation within the sporadic ALS/MND cohort. In the spinal cord, motor structures (ventral horns and corticospinal tracts) were most severely affected although the corticospinal tracts showed more marked inflammation than the ventral horns. Furthermore, there was involvement of extra motor regions (such as the spinothalamic tract) of the cord in many cases. This may be a pathological correlate of non-motor symptoms such as pain, which are increasingly recognised (23). The dorsal columns were the least affected but not completely spared.

The immunostaining confirmed spinal cord upregulation of ApoE and TYROBP (most pronounced in the corticospinal tracts) in sporadic ALS/MND, while higher TREM2 expression was associated with patients with longer survival times, suggesting a possible protective role for this pathway.

Data from project MINE were accessed to assess the influence of *APOE* genotype on MND inheritance. It was found that, in contrast to Alzheimer’s disease, where the ε4 haplotype is a risk factor, in MND, ε3 was a risk factor for MND, while ε2 and ε4 appeared to be underrepresented in the MND population.

### Inflammation in MND/ALS is greater in the spinal cord than the motor cortex

While the motor cortex had less inflammation than the spinal cord, it was not completely unaffected – there were changes in microglial morphology as well as upregulated TYROBP expression. Importantly, considerable inter-individual variation in the degree of inflammatory maker expression. This suggests there may be a small but variable inflammatory response that requires large sample sizes to be reliably detected by array technologies. This is consistent with the literature where a small post mortem study (n=11 sporadic ALS/MND and 9 controls) reported no evidence for motor cortex inflammation when false discovery rate correction was applied to array data, although there was some evidence of inflammation with ‘uncorrected’ p values (52). In contrast, a larger study of 31 sporadic ALS/MND and 10 controls found a significant upregulation of genes associated with immune pathways (4).

Of note, studies of sporadic disease contrast with a recent study of C9orf72-related MND, usually associated with more severe inflammation than sporadic disease (76), Using the same nCounter neuroinflammatory platform to assess the motor cortex of 10 MND-C9orf72 and 10 controls the authors observed that “the number of significantly dysregulated genes in this analysis is relatively low”. However, they found up regulation of a number of inflammatory markers that highlighted a variety of pathways (including NFκB, which was also highlighted in our own spinal cord data).

The differing intensity of MND-associated inflammation between motor cortex and spinal cord are not confined to MND: Ritzel et al. (78) found that mouse spinal cord and cortex had differing responses to ageing in terms of microglial activity and phagocytic potential as well as blood brain barrier integrity. Other studies have shown that spinal cord injury causes a greater microglial response and blood brain barrier breakdown than cortical injury (8, 9, 81). It is thus possible that spinal cord microglia have a lower threshold for activation than cortical microglia in a variety of pathological contexts. More human studies comparing spinal cord and cerebral inflammation are needed.

The motor cortex-spinal cord differences in inflammation may also be related to differing MND-related pathology: There is well-documented neuronal loss in the spinal cord (42). However, while neuronal loss has been described in the motor cortex (15, 64), more rigorous and stereological studies have shown no change in the number of motor neurons (33, 88).

Further, cystatin C proteinopathy in the form of Bunina Bodies is a pathological hallmark of MND, specifically present in the lower motor neurons and not a feature of motor cortex disease (10).

The finding of marked spinal cord inflammation fits well with a recent transcriptomic study that highlighted an increase of microglial and inflammatory markers in the spinal cord in a large cohort of ALS/MND cases that included both sporadic and familial cases (41).

### Disease-associated microglia and MND/ALS

We found upregulation of TREM2 mRNA in sporadic ALS/MND, with the protein levels associated with longer survival. This is in line with transcriptomic studies that have consistently found upregulated TREM2 in human MND, and have associated soluble TREM2 with neuroprotection (20, 24).

In the CSF, soluble TREM2 protein is highly expressed in the early stages of disease and diminishes with progression (24). In late-stage disease, soluble TREM2 expression positively correlates with survival time, suggesting this may be protective.

TREM2 is a cell surface receptor that regulates the inflammatory phenotype in myeloid cells (99). When activated by ApoE, the cytoplasmic domain complexes with TYRO protein tyrosine kinase-binding protein (TYROBP, also known as DAP12), which signals through an intracellular immunoreceptor tyrosine activation motif, which can result in an anti-inflammatory phenotype and phagocytosis (48). In support of an anti-inflammatory role for TREM2, reducing TREM2 expression and activity dampens the anti-inflammatory response following IL-4 stimulation in transgenic mouse microglia (56) and cultured microglia (100). In addition, TREM2 binds TDP-43 and TREM2 depletion in microglia causes loss of the ability to phagocytose TDP-43 inclusions, thereby enhancing motor dysfunction (97). An analogous phenomenon has been observed for amyloid ß animal models of Alzheimer’s disease (69), leading to early-stage trials of TREM2 agonism to treat Alzheimer’s disease (80). The possibility of using existing agents to treat MND/ALS is an exciting prospect.

TREM2 is widely expressed in the brain, and due to its expression by somatic human macrophages, and confirmed murine microglial expression, it has been assumed human microglia also express TREM2. However, while human microglia may express TREM2 mRNA (20, 102), immunohistochemistry studies have failed to find microglial TREM2 protein in human *post-mortem* brain. In contrast, there have been demonstrations of TREM2-positive cells in intravascular monocytes, neurons and perivascular macrophages (27, 73, 79). In the context of acute infarction, TREM2 is seen in cells in the brain parenchyma, possibly representing recruitment of monocytes under pathological conditions (27).

Similarly, we found TREM2 to label the same cells (intravascular and perivascular monocytes) as well as some neurons with few parenchymal cells expressing this protein in the motor cortex. There was a greater number of TREM2-positive parenchymal cells in the spinal cord and increased expression of spinal cord TREM2 in ALS/MND with longer survival. The TREM2-positive cells were also IBA1^+^ amoeboid cells, possibly representing recruited macrophages (27).

TYROBP expression in MND has not been as widely studied. We observed a significant upregulation of TYROBP mRNA in the spinal cord, as well as TYROBP protein in the motor regions of the spinal cord and in both the white and grey matter of the precentral gyrus.

The literature to date suggests a toxic role for TYROBP: Knockdown in a mouse model of hypoglossal nerve injury resulted in reduced proinflammatory cytokine production, and reduced neuron death (47). Similarly, reduced TYROBP function in mouse models confers resistance to demyelination (44) as well as tau hyperphosphorylation and dystrophic neurites in Alzheimer disease (36). Finally, TYROBP deficiency in mice seems to confer resilience to Alzheimer-type tau and amyloid ß pathology (36).

Upregulation of ApoE protein and *APOE* mRNA was detected in the spinal cord in MND consistent with previous studies (3, 66, 67). ApoE is a fat-binding protein involved in lipid transport, neuronal survival and plasticity, and neurite outgrowth (40, 50, 51, 92). In the brain, it is mostly expressed by astrocytes and microglia, with lesser expression by neurons (35, 51, 60, 71, 98).

Previous studies of CSF and serum in human MND showed evidence for defective clearance of excess toxic cholesterol forms and a deficit in related neuroprotective metabolites (1). It is possible that glial metabolic pathways are unable to cope with cholesterol release from dying neurons, resulting in increased toxicity and inflammation (11).

Extracellular ApoE protein can act as a ligand to the TREM2 receptor triggering microglial phagocytosis (6). Knockdown of *APOE* results in ineffective neuronal debris clearance in a model of prion pathology (68), implying a neuroprotective role for ApoE.

TREM2, TYROBP and ApoE together form a well-characterised pathway responsible for the Disease-Associated microglial (DAM) phenotype, which has a common signature across several models of neurodegeneration e.g. (39, 49, 84). Through activation of TREM2 signalling, often by ApoE, TYROBP results in the downregulation of Transforming Growth Factor β (TGFβ)-mediated microglial genes, and a simultaneous upregulation of the DAM genes (Figure Discussion 1). These DAM-associated markers regulate inflammation, lipid metabolism, phagocytosis and lysosomal pathways (26, 49).

DAM have been identified in many mouse models, particularly of AD, (39, 45, 49), but also including other neurodegenerative models such as ALS/MND (22, 39, 49, 84). *Post-mortem* studies have also found evidence of the DAM signature in AD (28, 46). However, while this DAM phenotype has been identified in models, this relevance is only emerging in human ALS/MND. Importantly, using the R Shiny app to interrogate transcriptomic data from a recent transcriptomic study (41) highlights findings that accord with our own, namely an upregulation of all three of *TREM2*, *TYROBP* and *APOE* in the spinal cord (*P*≤0.00057) in MND after false discovery rate correction. All three correlated negatively with disease survival in the cervical (*P*≤0.016) but not lumbar (*P*≥0.054).

### The APOE haplotype and amyotrophic lateral sclerosis/motor neuron disease

The *APOE* ε4 haplotype is one of the most important risk factors for the development of late onset AD (54). With respect to the relationship between MND and *APOE,* a 2014 meta-analysis of 4249 MND patients and 10,397 controls from North America, Scandinavia, Europe Israel and Guam have found no relationship (83). However, a Chinese study (n=683 MND patients and 369 controls) reported a modest association between MND and ε4 (Odds ratio 1.42; 95%CI, 1.02-1.98; p=0.02) (37). We addressed this question in the largest cohort analysed to date including 29,612 ALS/MND patients and 122,656 controls and found that MND patients were significantly enriched with the *APOE* ε3 haplotype but depleted of both the ε2 and ε4 haplotypes.

In addition, the ε2 haplotype in addition to being under-represented in the ALS/MND group was associated with a later age of onset. This apparent protection is supported by previous studies that have variously found the ε2 haplotype to be associated with a later age of onset (53, 72) or longer survival (63). The ε4 haplotype was also under-represented in the ALS/MND cohort, and was related to shorter survival, consistent with previous studies of ε4 favouring bulbar onset disease, which itself is associated with more severe disease (53, 63).

Collectively, our *APOE* data suggest that the different haplotypes are associated with different disease phenotypes with unique clinical presentations, risks and severity. This shift in *APOE* genotype in the ALS/MND population may be partially responsible for the altered expression levels seen (75).

### Limitations and future directions

This is the largest human *post-mortem* study of ALS/MND, providing a unique set of data. However, limitations of the study include that this is a snapshot of the end stage of disease, and as such, cannot assess the inflammatory status at earlier stages and cannot determine directions of causation. Nevertheless, the data support better experimental models.

The upregulation of members of the ApoE-TREM2-TYROBP pathway suggest this is an obvious candidate for therapeutic intervention as currently performed it already is in Alzheimer disease. However, the impact of increased expression remains to be examined in sporadic ALS/MND.

The relationship between *APOE* genotype and immune pathology is a key question. Unfortunately, this project was not resourced for this, and this will form the focus of future studies.

In conclusion, we have demonstrated marked and variable neuroinflammation in human sporadic MND-TDP that is most florid in the spinal cord, and significantly more subtle in the motor cortex and highlights the APOE-TREM2-TYROBP pathway in particular. Finally, we have performed the most high-powered study to date of the relationship between APOE genotype and sporadic ALS/MND and found that the ε2 and ε4 haplotypes appear protective, while the ε3 haplotype was a risk factor.

## Data Availability

All data produced in the present study are available upon reasonable request to the authors

## Acknowledgements

The authors thank the Pathological Society of Great Britain and the British Neuropathological Society for funding this project and to Bruker Spatial Biology who provided two of their nCounter Neuroinflammation panels. We are also grateful to the Sheffield Brain Tissue Bank for supplying the tissue and to those who have donated tissue for scientific research and their families who have supported this.

## Declarations

Author contributions (CRediT): Conceptualization, BAA, JES, DB, JCK, PRH, JRH; Data curation; BAA, JCK, PRH, WW, JRH; Formal analysis, BAA, JCK, WW, MD, JRH; Funding acquisition, JES, DB, PRH, JRH; Investigation, BAA, JES, CD, PRH, DF, CAM, JRH; Methodology, BAA, JES, CD, DB, JCK, PRH, DF, CAM, WW, MD, JRH; Project administration, BAA, JES, CD, DB, PRH, DF, JRH; Resources, JES, CD, PRH, DF, CAM, JRH; Software, BAA, WW, MD; Supervision, JES, PRH, JRH; Validation, BAA, CD, PRH, WW, MD; Visualisation, BAA, WW, JRH; Writing – original draft, BAA, JRH; Writing – review & editing, all authors. The majority of the data presented here formed the basis of a PhD project undertaken by BAA. The Sheffield Brain Tissue Bank (SBTB) which provided the tissue used here has ethical permission to function as a Research Tissue Bank. At the time the work was undertaken, this was covered by a favourable opinion from the Scotland A Research Ethics Committee (Reference 19/SS/0029). SBTB adheres to consenting protocols laid down by the UK Human Tissue Authority and agreed by Research Ethics Committee.

